# Customisable digital cognitive aids cut total error by 75% defining a threshold for irreducible human error in clinical care: a pooled analysis of five randomised trials

**DOI:** 10.1101/2025.10.04.25337246

**Authors:** Jean-Christophe Cejka, Clément Buléon, Marc Lilot, Antoine Duclos, François Lecomte, Karim Tazarourte, Baptiste Balanca, Thomas Rimmelé

## Abstract

**Background:** Human error is a leading cause of preventable harm in clinical care, driven by factors such as cognitive overload and staff turnover. While **customisable digital cognitive aids (cDCAs)** have emerged as real-time protocol-tailored support, a framework to quantify their total impact on human error and its fundamental limits has been lacking.

**Methods:** We conducted a pooled analysis of five randomised high-fidelity simulation trials with consistent methodologies, including 370 healthcare professionals across diverse clinical settings and levels of experience. Using bootstrap resampling, we modelled the distributions of technical and non-technical skills (TS and NTS) to quantify the impact of cDCAs on clinical competence and on **Total Human Error** (THE)—defined as the sum of systematic deviation from standards (bias^2^) and inter-individual variability (variance).

**Findings:** The use of **cDCAs reduced THE by 75%** in a novice user context. This effect was driven by concurrent, drastic reductions in systematic deviation from standards (**bias**^**2**^: 328.1 vs 1377.1) and inter-individual variability (**variance**: 124.3 vs 210.0). This was reflected in significant improvements in both technical (81.9±0.8 vs 62.9±1.0, p<0.001) and non-technical skills (84.9±0.8 vs 75.2±1.2, p<0.001), demonstrating enhanced clinical competence and robustness. Crucially, this substantial error mitigation revealed a consistent **residual error threshold**, quantifying an empirical upper bound of ∼25% for what we define as **Irreducible Human Error** (IHE).

**Interpretation:** By reducing both systematic bias and performance variability, cDCAs demonstrably harmonise clinical practices and enhance competence robustness. Our analysis provides the first empirical framework to quantify this beneficial effect, revealing in turn a ∼25% residual error that **delineates the fundamental limits of performance enhancement achievable** via procedural workflow support. We posit this threshold serves as an empirical upper bound for Irreducible Human Error (IHE)—errors arising not from procedural flaws but from higher-order cognitive or interpersonal factors. Establishing IHE as a new benchmark for patient safety thus provides foundational evidence for a model of augmented practice, where complementary innovations such as AI may be required to move beyond procedural support, augment clinical judgement, and uphold human-centred care.

**Research in Context:** *Evidence before this study:*

- Cognitive support has long been used to mitigate human error in high-stakes environments.
- ‘Traditional’ cognitive support tools (e.g., paper checklists) improve adherence to technical protocols during medical crises, but their impact on non-technical skills—such as communication and decision-making—is limited and their clinical adoption remains low.
- Early digital aids largely replicated these static tools, often lacking the ergonomic integration needed for real-time clinical use and failing to leverage the full potential of digital platforms.
- Consequently, no prior study had systematically assessed the impact of **customisable** digital cognitive aids (cDCAs) on both technical and non-technical skills across diverse clinical domains to establish their overall **efficiency** and **robustness**.
- Furthermore, no prior study has established a comprehensive framework for evaluating their effects on ‘Total Human Error’ (THE)—encompassing both bias and variability in practice—or introduced a measurable threshold for an ‘Irreducible Human Error’ (IHE) in clinical care.

*What this study adds:*

- This study is the first to quantify competence—defined as the integration of procedural proficiency (TS) and cognitive-behavioural dimensions (NTS)—and measured the impact of cDCAs on TS and NTS performance and variability, across multiple clinical settings and experience levels. We pooled five randomised controlled trials with consistent methodologies and used a resampling technique (bootstrap analysis), which allowed us to model distributions of performance and assess robustness across diverse professional backgrounds.
- Unlike traditional digital checklists, which often impose cognitive effort or disrupt team workflows, cDCAs are adaptive interfaces, integrating seamlessly into decision-making with minimal cognitive load.
- Our work systematically quantifies clinical competence and THE across both TS and NTS. We demonstrated that cDCAs significantly reduced systematic bias—defined as deviation from expected standards—along with inter-individual and inter-situation variability, thereby harmonising clinical practices and improving reliability—even in high-stakes scenarios. The reduction in inter-situation variability is particularly relevant, as it strengthens the robustness of clinical care delivery, ensuring consistent performance across diverse settings.
- By defining and quantifying IHE, we establish a benchmark for understanding the limits of human performance—beyond which cognitive aids alone may be insufficient.

*Implications of all the available evidence:*

- Integrating customisable, ergonomically optimised cDCAs into routine clinical workflows provides a scalable solution to reduce THE by harmonising practices and improving adherence to care standards. This is particularly valuable in resource-limited settings, where cDCAs could help exchange expertise.
- By establishing IHE as a measurable threshold, this study provides a foundation for further innovations in cognitive support. While current cDCAs provide substantial reductions in THE, these findings raise the question of whether AI could further complement cognitive aids while preserving human expertise at the core of decision-making.
- Future research should explore how AI-enhanced cognitive aids might ethically and transparently address residual human error, ensuring these technologies reinforce—rather than undermine—the foundational principles of trust, accountability, and patient-centred care.
- Finally, beyond individual performance, cDCAs may contribute to global equity by promoting robust, harmonised care and enhancing knowledge retention and application across healthcare systems. In an interconnected world, where no system operates in isolation, ensuring safe, high-quality care on a global scale is imperative.

## Introduction

Human error, driven by the relentless pressures of cognitive overload and high staff turnover in modern healthcare, remains a leading cause of preventable harm [1–4]. While medicine has long looked to high-reliability industries for solutions, the adoption of first-generation tools like paper checklists proved to be a partial remedy[5–7]. These static aids improved adherence to technical protocols but largely failed to enhance the crucial non-technical skills that underpin team performance in a crisis, leaving the culture of patient safety fundamentally unchanged. A new generation of customisable digital cognitive aids (cDCAs) have emerged as a more potent solution[8,9]. Functioning as ‘cognitive interfaces,’ these tools restructure action by offloading low-value cognitive tasks—such as protocol adherence, time-keeping, and calculations—thereby freeing healthcare professionals to focus on higher-value activities: dynamic situational assessment, complex decision-making, and patient-centred communication. Multiple randomised trials have demonstrated that this approach significantly enhances both technical and non-technical skills (TS and NTS) [10–14], and doubles knowledge retention[15]. This proven capacity to improve performance across varied clinical settings and experience levels underpins what we define as **robust clinical competence**: the consistent and reliable integration of skills under disruption[16,17].

This established ability of cDCAs to foster robust competence provides a stable foundation from which to address a more fundamental question: beyond improving performance, what is their ultimate impact on the very structure of human error? This requires moving beyond simple observation to model its underlying architecture. To do so, we introduce a framework to quantify **Total Human Error (THE)** as the sum of its two core components: systematic deviation from standards (bias^2^) and inter-individual performance variability (variance). These components represent the objective, quantifiable facets of error that structured cognitive support is designed to mitigate.

This framework, however, forces a confrontation with a critical question at the frontier of human performance: what remains when these errors are minimised? We hypothesised the existence of a residual threshold of Irreducible Human Error (IHE), composed of errors rooted in dynamic interpersonal and higher-order cognitive factors that cannot be overcome by procedural support alone. Defining this threshold creates a new boundary for patient safety and raises fundamental questions about how complementary innovations, such as artificial intelligence (AI), might be required to address this final fraction of error, augment clinical judgement and uphold human-centred care.

Here, we provide the first empirical test of this framework. In a pooled analysis of five randomised trials, we use bootstrap resampling to model performance distributions and rigorously quantify the impact of cDCAs on clinical competence, its robustness, and Total Human Error. This work also provides the foundational evidence for a new model of augmented clinical practice, framing cDCAs as a powerful cognitive interface that bridges the gap between knowledge and competent action.

Our primary objective was to calculate the magnitude of THE reduction and to provide the first empirical estimate of the Irreducible Human Error threshold. By seamlessly supporting decision-making within integrated workflows cDCAs pave the way for future synergies

## Materials and methods

### Study design

Data were pooled from five randomised controlled trials (RCTs) conducted at our institution[10–14] using consistent methodologies including remote double evaluation by expert simulation instructors, and previously published in peer-reviewed journals. These studies assessed the impact of customisable digital ‘cognitive aids’ (cDCAs), created and improved by our university team (MAX by MEDAE, Lyon, France, https://www.medae.co/en/ since 2015, to manage high-stakes situations within high-fidelity simulated environments. The cDCAs were tested across diverse clinical and paramedical settings, spanning specialities from anaesthesiology and military medicine to obstetrics and neonatology. In all scenarios, the team leader either had access to the cDCAs or managed the situation without it. Overall, the analysis included 370 participants, comprising 186 who used the cDCAs and 184 who did not.

Technical skills performance was evaluated using scores adapted from international standards of care, while NTS were evaluated using validated scales[19,20]. Scores were normalised from 0 to 100, representing percentages of the maximum achievable score. Mean Squared Error (MSE = bias^2^ + variance) served as an estimator of ‘Total Human Error’ (THE).

### Data resampling and distribution modelling

The dataset was analysed using bootstrap resampling (20,000 bootstrap iterations per parameter), a robust method for generating distributions of key parameters, including mean, variance, bias^2^, and Mean Squared Error (MSE).

We opted for bootstrap resampling rather than a traditional meta-analysis, as all included RCTs followed a standardised methodology with homogeneous assessment tools and simulation protocols. Unlike meta-analyses, which typically aggregate point estimates, the bootstrap approach was chosen specifically for its ability to model full empirical performance distributions. This **non-parametric, frequentist method allows for a robust estimation of variance and higher-order moments without assuming a prior probability distribution** for the parameters of interest.

The MSE was decomposed into its two components, Bias^2^— here defined as the squared deviation from an ideal performance score of 100, a metric of systematic inaccuracy—and Variance—capturing the spread of performance outcomes, highlighting inter-individual variability in caregiving practices. This analysis offered detailed insights into performance consistency and THE between cDCAs users and non-users.

In addition, the variance distributions of each bootstrap sample were examined. The mean of these distributions represents the overall performance variance, while their spread (variance of variances) provides deeper insight into performance variability across different professional backgrounds and clinical environments. This metric is essential for evaluating the robustness of care delivery with cDCAs in diverse conditions.

Further methodological details, including the bootstrapping procedure and statistical considerations, are provided in **Supplementary Appendix 1**.

### Statistical Analysis

The normality of the distributions of means and variances obtained from the bootstrap analysis was assessed using the Shapiro-Wilk test. We applied parametric Welch t-tests, which do not assume equal variances, to evaluate statistical differences between the groups. To compare the variances of the resampled populations, the Levene test was employed.

## Results

### Mean performances and inter-individual variability

Significant improvements were observed in both TS and NTS with the use of cDCAs. Mean scores were higher in the cDCAs group compared to the No-cDCA group (TS: 81.90±0.81 vs 62.93±1.05, p<0.001; NTS: 84.9±0.8 vs 75.2±1.2, p<0.001), with corresponding decreases in variances (p<0.001, Fig. 1).

**Figure 1:**
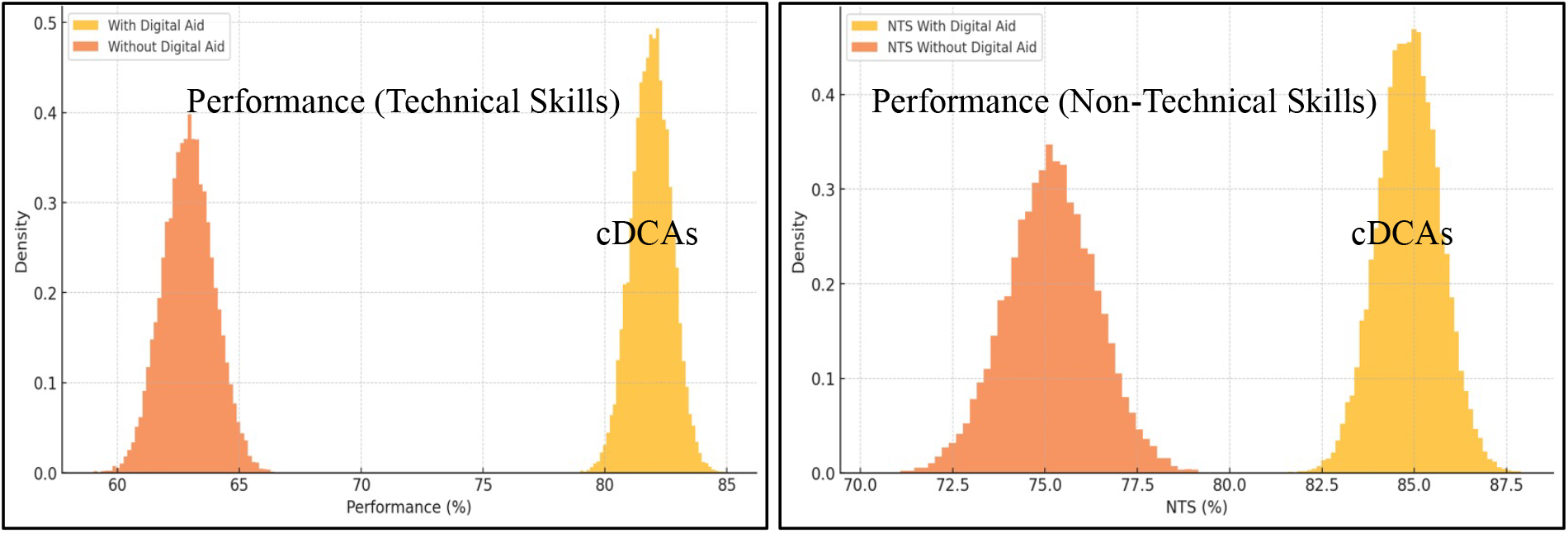
Bootstrap distributions of Technical (left panel) and non-technical skills (right panel) performance with cDCAs (in yellow) and without cDCAs (in orange).

Bias^2^ distributions, reflecting deviations from the ideal score of 100%, were significantly lower in the cDCAs group (TS: 328.1±29.2 vs 1377.1±78.8, p<0.001; NTS: 230.0±25.5 vs 618.1±60.9, p<0.001). Variance in bias^2^ was also significantly reduced (TS and NTS: p<0.001), indicating improved accuracy and reduced systematic errors with cDCAs.

### Inter-situation variability

The resampled variance distributions for the cDCAs group were centred on significantly lower values compared to the No-cDCA group (TS: 124.3±11.2 vs 210.0±19.6, p<0.001; NTS: 126.9±11.1 vs 267.2±25.7, p<0.001, Fig. 2). This suggests more consistent performance and reduced inter-situation variability in the cDCAs group, reflecting harmonised care delivery across diverse scenarios.

**Figure 2:**
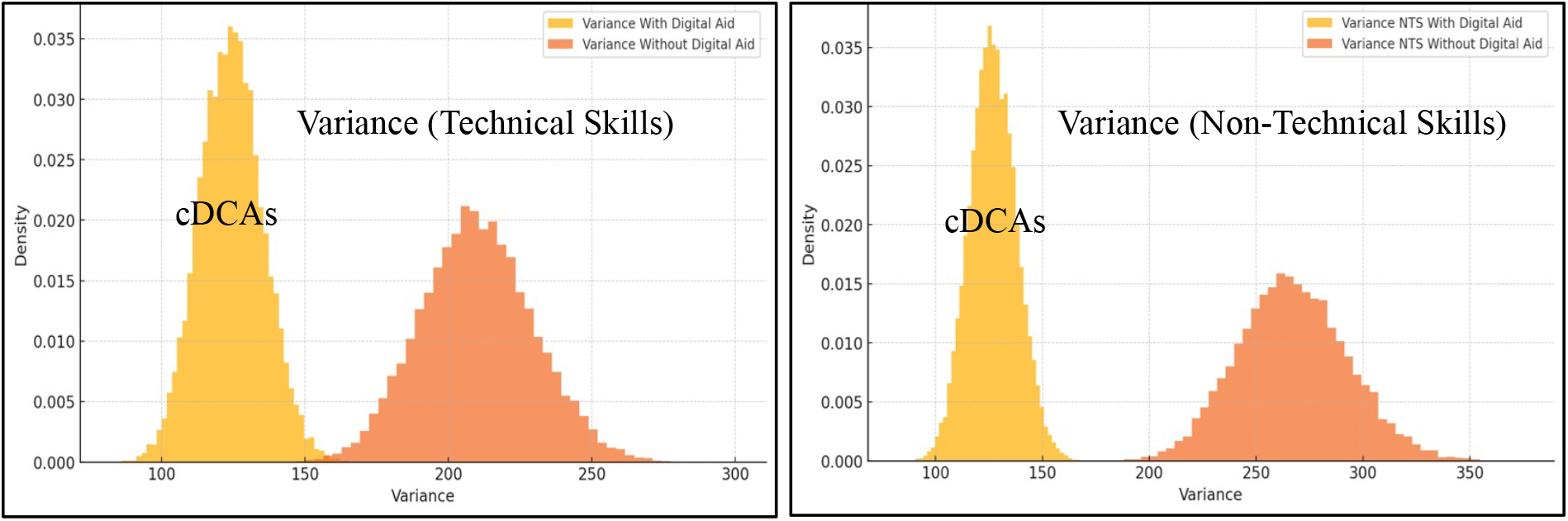
Bootstrap distributions of the variance for Technical Skills (left panel) and Non-Technical Skills (right panel), with cDCAs (in yellow) and without cDCAs (in orange).

The spread of the variance distributions (variance of variances) was also significantly smaller in the cDCAs group (TS and NTS: p<0.001), demonstrating greater robustness and uniformity in care delivery across professional backgrounds and clinical care environments.

### Mean Squared Error (MSE)

The use of cDCAs was associated with a marked reduction in Mean Squared Error (MSE), reflecting decreased THE. MSE values were significantly lower in the cDCAs group (TS: 452.3±36.1 vs 1587.1±82.9, p<0.001; NTS: 356.9±31.2 vs 885.3±71.9, p<0.001, Fig. 3). The variance of MSE was also significantly reduced, further supporting the consistent effectiveness of cDCAs in improving performance accuracy and reducing variability.

**Figure 3:**
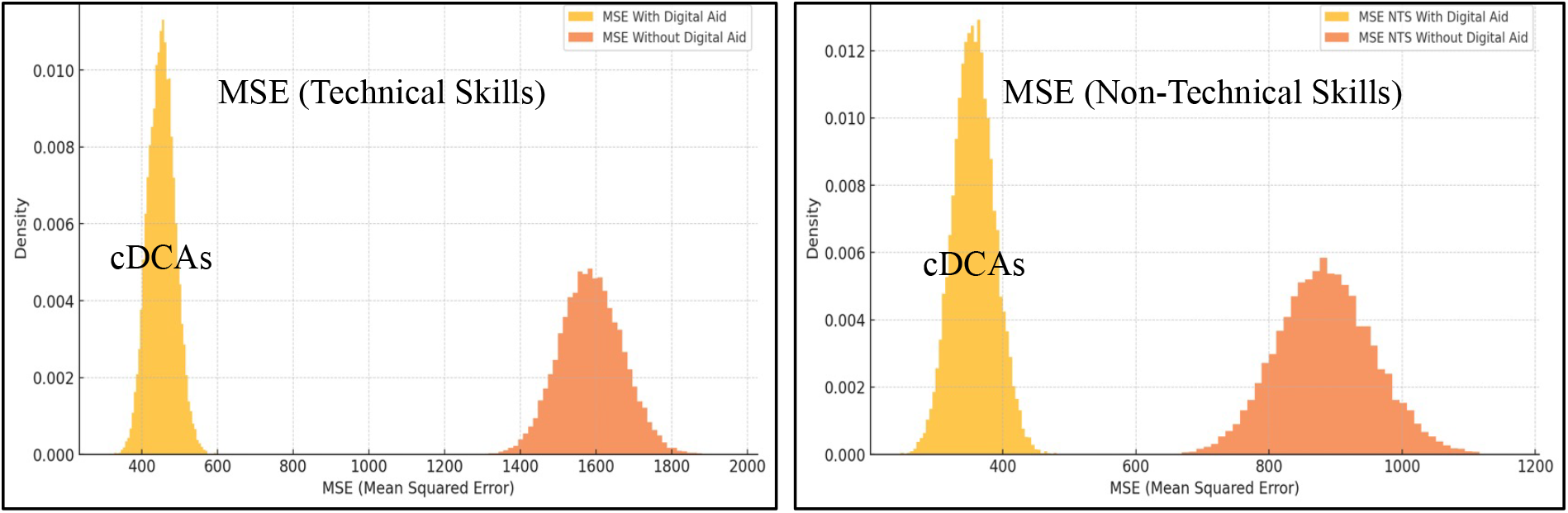
Bootstrap distributions of Total Human Error, estimated by the Mean Squared Error (MSE = bias^2^+variance) for Technical Skills (left panel) and Non-Technical Skills (right panel), with cDCAs (in yellow) and without cDCAs (in orange).

## Discussion

### Enhancing Clinical care Performance through Digital Cognitive Aids

Our findings establish that customisable digital cognitive aids (cDCAs) are pivotal innovations for enhancing clinical competence. This pooled analysis of five randomised trials provides the first large-scale, quantitative evidence that cDCAs systematically reduce both bias and inter-individual variability—the core components of Total Human Error—thereby fostering a more robust and standardised model of care. Building on foundational research into medical error[1–4] and cognitive support[9], this work demonstrates a paradigm shift in patient safety, moving from simple task adherence to the measurable enhancement of robust clinical competence.

The mechanism for this enhancement lies in cognitive offloading. By automating routine, low-yield cognitive tasks—such as protocol adherence, calculations, and timekeeping— cDCAs alleviate the clinician’s cognitive load. This frees them to focus on high-value tasks like team coordination, situational reassessment, and patient-centred care, aligning with cognitive science principles on the delegation of routine tasks, enabling professionals to deal with complex and unpredictable aspects of patient care requiring intuition and expertise. This directly supports the development of **competence**, a construct integrating not just knowledge and practical skills, but also professional behaviours, as outlined in frameworks like CanMEDS[17]. By supporting professionals even in challenging conditions, cDCAs lower **Total Human Error** (THE), an approach that echoes the safety culture in high-stakes fields such as aviation and nuclear safety.

Furthermore, by reducing inter-situation variability—as evidenced by the decreased variance of variances in our bootstrap analysis—cDCAs contribute directly to clinical care **robustness**: the ability of a system to maintain its functional integrity under disruption[16]. This involves ensuring reliable, high-quality care despite varying conditions, such as shifts in team expertise or patient complexity. These findings indicate that broader adoption of cDCAs in both training and clinical teamwork is not only feasible but highly impactful, representing a scalable paradigm shift in patient safety, particularly in resource-limited settings.

### Quantifying Irreducible Human Error: A New Benchmark for Safety

With minimal prior training (∼15 minutes per participant) across the five randomised trials, cDCAs substantially reduced THE, revealing in turn a residual error. We posit that this error—a composite of persistent cognitive and interpersonal factors— delineates an **empirical upper bound** for what we define as ‘**Irreducible Human Error’** (IHE). Our analysis reveals this remainder is primarily driven by residual bias (residual bias^2^: 328.1 vs 1377.1), though inter-personal variability (variance: 124.3 vs 210.0) also contributes significantly, a finding consistent with analyses of human error in many professional domains[21].

This threshold of ∼25% within a novice user context does not necessarily represent an absolute physiological or cognitive limit. Real-world implementation of cDCAs suggests that this residual error can be further compressed. Teams using the cDCA as a routine cognitive interface in different clinical settings—ranging from anaesthesiology-intensive care to routine delivery room management— consistently achieve 90–95% of expected technical performance, even in overwhelming situations—provided the appropriate procedure is accessed within the cDCAs, diagnosis errors being counted into variance. This extensive field experience leads us to hypothesise that the **asymptotic IHE** for practiced users **may converge towards a 10–15% threshold**. This figure represents a prospective benchmark for optimising cognitive support systems and **defines the frontier where complementary support strategies may be required**.

However, this purely quantitative threshold prompts a deeper, qualitative question: what does ‘perfect performance’ truly mean in the complex reality of clinical care? Beyond numerical performance scores, the concept of ‘good care’ remains inherently complex. While performance metrics provide valuable insights into technical execution, they fail to fully capture the nuances of clinical decision-making and patient outcomes. For instance, in critical scenarios such as maternal cardiac arrest, administering an incorrect adrenaline dose or failing to displace the uterus leftward may both be recorded as single missed items, yet they carry vastly different prognostic implications. Thus, striving for 100% performance scores with cDCAs does not inherently guarantee better outcomes, but it offers two key advantages. First, it maximises patient safety by ensuring that no high-stakes actions—such as uterine displacement in maternal cardiac arrest—are overlooked. Second, it provides psychological reassurance to caregivers, allowing them to dedicate their cognitive resources to high-value tasks like team coordination and situational reassessment.

Over several years of field experience with cDCAs, team leaders have consistently reported a ‘sense of accomplishment’, confident that they ‘did everything possible’ during critical events. This confidence directly translates into more effective post-event debriefings, fostering continuous team improvement and encouraging broader adoption of cDCAs in routine clinical practice.

### Future Directions: Augmenting Cognition with Artificial Intelligence

The persistence of IHE, even when minimised, underscores the need for complementary innovations. Clinical care decision-making is dynamic and requires tools that can manage large volumes of data while adapting to evolving conditions. Through the lens of Shannon’s communication theory[22], current cDCAs already contribute to reducing entropy—quantified uncertainty—by standardising guidance, minimising variability, and facilitating the continuous reassessment of hypotheses[23].

One promising avenue is the integration of artificial intelligence (AI), which could provide real-time hypothesis generation and offer context-aware decision support, thus reducing the cognitive load required for data synthesis in complex cases. Yet, AI carries inherent challenges, including algorithmic opacity (‘black-box’ decision-making), bias in training data, ethical and medico-legal concerns, and potential over-reliance on automated systems.

While AI can provide timely insights, core human attributes—empathy, accountability, teamwork, and ethical judgement—remain irreplaceable. Designing AI-enhanced cognitive aids that complement human expertise rather than seeking to replace it, ensures that these tools remain aligned with the patient-centred values at the core of clinical care delivery.

This vision of AI-enhanced cognition resonates with Alan Turing’s seminal reflections on ‘machine’ intelligence[24]. As early as 1951, Turing envisioned systems capable of learning from mistakes and improving autonomously over time. As he presciently cautioned, such systems could also introduce unexpected errors, deterministic but opaque, that defy human intuition. Turing’s reflection, a cornerstone in the early development of artificial intelligence, is particularly relevant today in clinical practice, in which challenges such as algorithmic opacity, biases in training data, and generalisation errors could undermine trust in AI systems. These errors, pose unique challenges that require careful oversight to mitigate, and AI algorithms, while powerful, should never be regarded as infallible or treated as ‘divine truth.’ Their application requires constant critical scrutiny to ensure that algorithmic errors do not translate into errors in patient care.

While these perspectives on AI are promising, our findings reaffirm the critical and immediate role of cDCAs in addressing current clinical care challenges. Unlike speculative AI applications, cDCAs have already demonstrated their capacity to standardise practices, reduce variability, and support robust care delivery across diverse settings. The COVID-19 pandemic has further underscored the urgent need for customizable, scalable, equitable solutions to strengthen global healthcare systems. By bridging gaps in practice and enabling consistent, high-quality care, cDCAs represent an operational response to these challenges, particularly in resource-limited environments. Equitable access to these technologies is not only an ethical imperative but also a practical necessity for improving clinical care outcomes worldwide.

### Limitations of the study

This study is subject to several limitations. First, it was conducted exclusively in simulated environments. Although simulation-based research is widely recognised for its predictive value in real-world clinical care settings, the results may not fully capture the intricacies and unpredictability of live clinical care. Nonetheless, the homogeneity of simulation methods across the included trials ensured consistency and comparability, which was critical for the study’s primary objective of modelling performance distributions. This methodological coherence differentiates this study from a meta-analysis, as the pooled data originated from trials employing the same cDCAs and evaluation criteria.

Second, while bootstrap resampling offers a robust statistical framework for modelling distributions and estimating variance, its accuracy depends on the quality and representativeness of the original dataset. In this study, the dataset was consistent across trials, as all were conducted within the same institutional research network using standardised simulation protocols. To ensure the robustness of the bootstrap-generated distributions, convergence tests were performed with varying numbers of iterations. No further improvements in precision were observed beyond the chosen threshold of 20,000 iterations, strengthening confidence in the coherence and reliability of the modelled distributions.

## Conclusion

In conclusion, our findings confirm that cDCAs drastically reduce Total Human Error and significantly improve clinical robustness. However, the very act of quantifying this ∼75% reduction forces us to confront two fundamental challenges that define the future of patient safety. First, how might we **address the residual Irreducible Human Error** with more advanced tools, such as artificial intelligence? Second, and more fundamentally, how can we **ensure** that the potential demonstrated here is realised in practice, given the historical failure of **cognitive tool adoption** in medicine?

These two questions form the critical next steps in building a truly augmented model of clinical competence.

## Data Availability

All data produced in the present study are available upon reasonable request to the authors

## Author contributions (CRediT)

Conceptualization: JCC

Methodology: AD, BB, CB, JCC

Formal Analysis: BB, CB, FL, JCC, ML

Investigation: JCC

Data Curation: All authors

Writing – Original Draft: JCC

Writing – Review & Editing: All authors

Visualization: JCC

Supervision: BB, CB, JCC, ML, TR.

## Supplementary Appendix 1: Bootstrapping methodology

The original dataset distribution was estimated using bootstrap resampling. Efron[18] introduced this technique for estimating the distribution of a statistical estimator by resampling with replacement from the original dataset. Essentially, the bootstrap method involves drawing repeated samples, typically mirroring the size of the original dataset, directly from the dataset itself. Each sample’s statistic is recalculated, producing an empirical distribution that sheds light on the statistic’s variability. This approach facilitates the estimation of standard errors, confidence intervals, and other indicators of statistical precision, providing deeper insights into the data’s underlying patterns. The bootstrap method provides a flexible, non-parametric approach to statistical inference, circumventing assumptions about the underlying distribution that parametric methods often require. Moreover, bootstrap techniques are invaluable for assessing the stability and confidence of estimators beyond the capabilities of classical inference methods.

All calculations and figures were performed with Python (V3.12.3). Twenty thousand (20,000) bootstrap iterations were executed on the pooled dataset to generate distributions of the mean, variance, bias^2^, and MSE for each group. The choice of 20,000 iterations balances precision and practicality, ensuring statistical reliability and robustness as supported by both empirical evidence and theoretical frameworks.

For each iteration, a mean and a variance value were calculated, enabling the sampling of distributions for each parameter of interest. These distributions were used to analyse:

- **Distributions of means**: Reflecting central values and inter-individual variability for each group.
- **Distributions of variances**: Highlighting inter-situation variability and the consistency of performance outcomes.

Finally, MSE was calculated using the following formula:

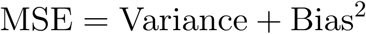

Where:

- Variance measures the dispersion in caregiving practices across participants.
- Bias^2^ represents systematic deviations from ideal performance (100%).

This decomposition allowed for a comprehensive understanding of how cDCAs influence Total Human Error and variability in clinical performance.

